# ETDRS letter scoring method improves the accuracy of 1.25% low-contrast visual acuity measurement in optic neuritis secondary to MS

**DOI:** 10.1101/2022.08.24.22279101

**Authors:** Yuyi You, Peng Yan, Stuart L. Graham, Alexander Klistorner

## Abstract

**Objectives:** To examine whether the dynamic range of 1.25% low-contrast visual acuity (LCVA) measurement in MS patients with optic neuritis (ON) can be improved by using the Early Treatment Diabetic Retinopathy Study (ETDRS) letter scoring method.

**Methods:** LCVA was tested using 2.5% and 1.25% low contrast ETDRS-style 4m Sloan letter charts. When ≥20 letters were read correctly, the letter score was equal to letter count plus 30. If <20 letters were read correctly, the letter score was equal to letter count at 4m plus the total number of letters read correctly at 1m.

**Results:** 51 relapsing-remitting MS patients with unilateral ON were enrolled and 60.8% ON eyes had a 1.25% LCVA letter count worse than 1 line (5 letters). In ON eyes with <20 letter count, ETDRS letter score showed a significantly improved correlation with both macular GCIPL thickness (r=0.71, p<0.0001; vs r=0.31, p=0.08 for letter count) and VEP latency (r=-0.47, p=0.003; vs r=-0.15, p=0.4 for letter count).

**Discussion:** Given ON with VEP monitoring has been frequently used as a model in MS remyelination clinical trials, our proposed ETDRS-style LCVA letter scoring method may be considered to enhance the functional outcome measure, which is necessary for regulatory approval.

---

Low-contrast visual acuity (LCVA) has been widely used as a primary outcome measure in multiple sclerosis (MS) studies and clinical trials.^1^ LCVA at 1.25% contrast level is often poor in optic neuritis (ON) eyes with a letter count close to 0, which may affect the accuracy of its measurement and its application in assessing axonal loss or de/remyelination in the disease process. In studies on retinal diseases, where patients usually have impaired central vision, the Early Treatment Diabetic Retinopathy Study (ETDRS) visual acuity is first measured at 4m and at 1m for patients with reduced vision.^2-4^ Given the LCVA charts with Sloan letters used in MS research are identical to the high-contrast ETDRS charts except that the letters are of lower contrast levels, we aim to determine whether the 1.25% LCVA can be scored using a letter scoring method similar to that being used in visual acuity testing with high-contrast ETDRS charts.

## Methods

### Standard protocol approvals, registrations, and patient consents

Consecutive relapsing-remitting MS patients with a history of unilateral ON were included in this study. The study cohort has been described previously in other studies.^5^ The study adhered to the tenets of the Declaration of Helsinki and was approved by the University Human Research Ethics Committee. Written consent was signed by all participants.

### LCVA measurement and statistics

LCVA was measured using 2.5% and 1.25% low contrast charts (4m/13ft Sloan letter logarithmic translucent contrast charts with No. 2425 illuminator cabinet, Precision Vision, La Salle, USA). For the letter score calculation, when ≥20 letters were read correctly at 4 m, the letter score was the total number of letters read correctly plus 30. If <20 letters were read correctly at 4m, the letter score was equal to the total number of letters read correctly at 4m plus the number of letters read correctly at 1m (only the 30 letters on the top 6 lines).^4^ The LCVA letter score was then correlated with macular ganglion cell inner plexiform layer (GCIPL) thickness and VEP latency (previously reported) using Spearman’s correlation (GraphPad Prism).

## Data Availability

The authors confirm that the data supporting the findings of this study are available within the article and from the corresponding author on reasonable request.

## Results

51 consecutive relapsing-remitting MS patients were included in this study and the demographic data of the study subjects have been described previously.^5^ For 2.5% LCVA, majority of the eyes had a better letter count than 20 (Table 1), therefore in this scenario the ETDRS letter score would be linearly correlated with the letter count (+30). However, for 1.25% LCVA testing, less than 1/3 of the ON eyes can read more than 20 letters which led to a significant disproportion between ETDRS letter score and simple letter count. Moreover, 60.8% ON eyes had a letter count worse than 1 line (5 letters) at 4m which may negatively affect the accuracy of LCVA data quantification. When the new scoring method was used, Spearman r between 1.25% LCVA and GCIPL improved from 0.50 (p = 0.001) to 0.72 (p < 0.0001). A secondary analysis was carried out in ON eyes with <20 letter counts and the improved accuracy of 1.25% LCVA measurement became more evident, with Spearman r increased from 0.31 (p = 0.08) to 0.71 (p < 0.0001) (Figure 1).

**Table 1:** Proportion of eyes that can read ≥20 letters in LCVA testing.

| Contrast | ON | fellow | P values |
| --- | --- | --- | --- |
| 2.5% | 74.5% | 96.1% | 0.004 |
| 1.25% | 31.4% | 58.8% | 0.009 |

**Figure 1:**
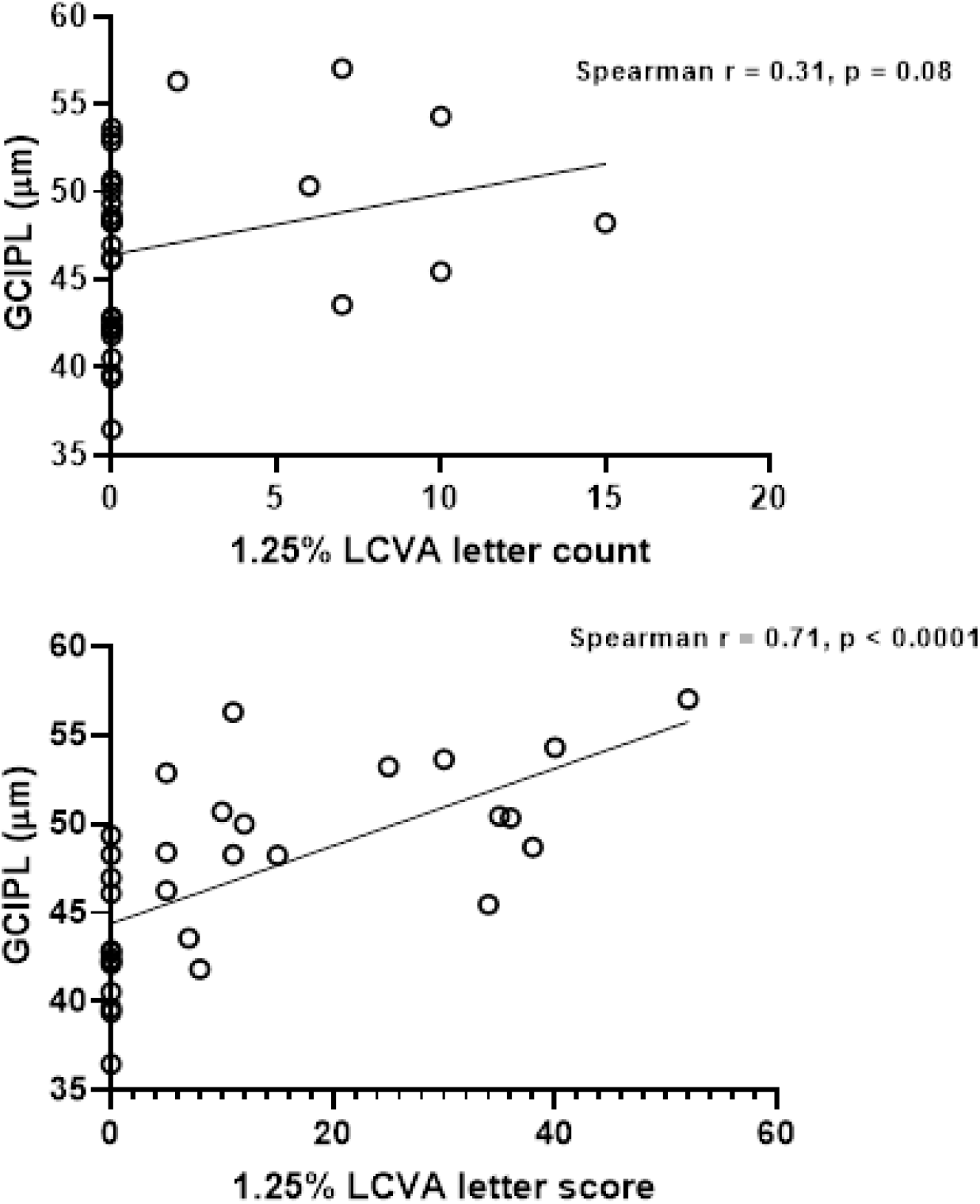
Improved correlation between GCIPL and 1.25% LCVA letter score compared to letter count in ON eyes with <20 letter count.

In addition, we found an increased correlation between LCVA and VEP latency in ON eyes using this new LCVA scoring approach (r = -0.44, p = 0.001 for letter score and r = -0.26, p = 0.07 for letter count). Again, in ON eyes with poor LCVA (<20), the correlation greatly improved between 1.25% LCVA and VEP latency (r = -0.47, p = 0.003 for letter score and r = -0.15, p = 0.4 for letter count, Figure 2).

**Figure 2:**
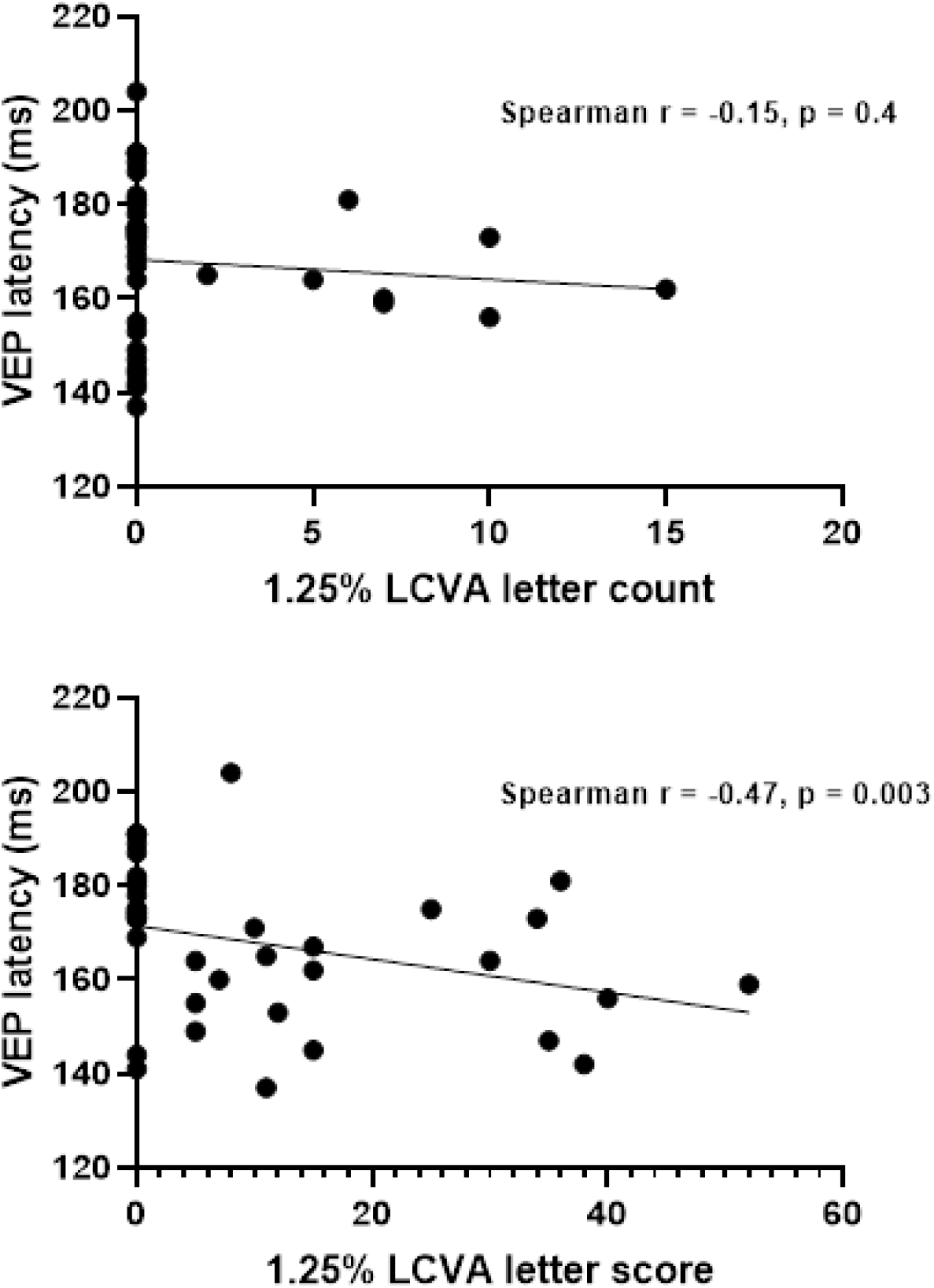
Improved correlation between VEP latency and 1.25% LCVA letter score compared to letter count in ON eyes with <20 letter count.

## Conclusions

This study shows that in ON eyes the ETDRS-style letter scoring method can improve the accuracy of 1.25% LCVA measurement, providing better correlation with both neuroaxonal loss (GCIPL) and demyelination (VEP latency) in ON eyes. It is important to note that the overall correlation between LCVA and GCIPL was stronger than that between LCVA and VEP latency, which supports our previous finding that LCVA reflects demyelination only in ON eyes with mild retinal ganglion cell loss.^6^ Given ON with VEP monitoring has been widely used as a model in MS remyelination clinical trials^7^, our proposed ETDRS-style 1.25% LCVA testing approach may be implemented to enhance the functional outcome measure, which is necessary for regulatory approval. In cases where 2m LCVA charts are used^1^, the charts can be moved from 2m to 0.5m (4:1 ratio) based on the constant geometrical progression of optotype size between lines.^3^

## Data Availability

All data produced in the present study are available upon reasonable request to the authors.

